# Durability of anti-Spike antibodies in the infant after maternal COVID-19 vaccination

**DOI:** 10.1101/2021.11.17.21266415

**Authors:** Lydia L. Shook, Caroline G. Atyeo, Lael M. Yonker, Alessio Fasano, Kathryn J. Gray, Galit Alter, Andrea G. Edlow

**Affiliations:** Massachusetts General Hospital, Department of Obstetrics & Gynecology; Ragon Institute of MGH, MIT, and Harvard; Massachusetts General Hospital, Department of Pediatrics; Brigham and Women’s Hospital, Department of Obstetrics & Gynecology

## Abstract

COVID-19 vaccination in pregnancy generates functional anti-Spike IgG antibodies that are known to cross the placenta. However, the durability of vaccine-induced maternal anti-S IgG in infant circulation, and how it compares to durability of antibody from maternal natural infection, is unknown. We quantified anti-S IgG in 92 2-month and 6-month-old infants whose mothers were vaccinated in pregnancy, and in 12 6-month-old infants after maternal natural infection with SARS-CoV-2. In the vaccinated group, 94% (58/62) of infants had detectable anti-S IgG at 2 months, and 60% (18/30) had detectable antibody at 6 months. In contrast, 8% (1/12) of infants born to women infected with SARS-CoV-2 in pregnancy had detectable anti-S IgG at the 6-month timepoint. Vaccination resulted in significantly higher maternal and cord titers at delivery and significantly greater antibody persistence in infants at 6 months, compared to natural infection.

## Introduction

COVID-19 vaccination in pregnancy generates functional anti-Spike (S) IgG antibodies in the maternal circulation that are detectable in the umbilical cord blood at birth.^1^–^4^ Anti-S IgG titers in the umbilical cord at birth are correlated with maternal titers and are highest after late second and early third trimester vaccination, although cord titers after first trimester vaccination have not yet been examined.^2^–^4^ The persistence of vaccine-induced maternal anti-S IgG in infant blood has not yet been characterized, nor compared to persistence of infant anti-S IgG after maternal natural infection.

## Methods

90 pregnant women who received one of three available COVID-19 vaccines in pregnancy and 12 pregnant women infected with SARS-CoV-2 in pregnancy who had enrolled in a prospective study at two large academic medical centers in Boston, MA enrolled their infants (n=104) in this follow-up study. Matched maternal and umbilical cord sera were collected at birth. Infant capillary sera were collected via microneedle device at either 2 months (n=62 vaccination) or 6 months (n=30 vaccination,12 infection) after birth. The study was approved by the IRB and all participants provided written informed consent. Antibody titers against the SARS-CoV-2 Spike protein were quantified using an enzyme-linked immunosorbent assay (Supplemental Methods).

## Results

Study participant characteristics are detailed in the Table. In the vaccinated group, 94% (58/62) of infants had detectable anti-S IgG at 2 months, and 60% (18/30) had detectable antibody at 6 months (Figure 1A-B, Table). In contrast, 8% (1/12) of infants born to women infected with SARS-CoV-2 in pregnancy had detectable anti-S IgG at the 6-month timepoint. Vaccination resulted in significantly higher maternal and cord titers at delivery and significantly greater antibody persistence at 6 months, compared to natural infection (Figure 1A-B, Table). Among vaccinated individuals, infant anti-S titers at 2 months were strongly correlated with both maternal and cord titers at delivery (Figure 1C). In contrast, neither maternal nor cord titers were strongly correlated with infant anti-S titers at 6 months, largely due to 40% of infants having no detectable titer at that time (Figure 1D).

**Table.**
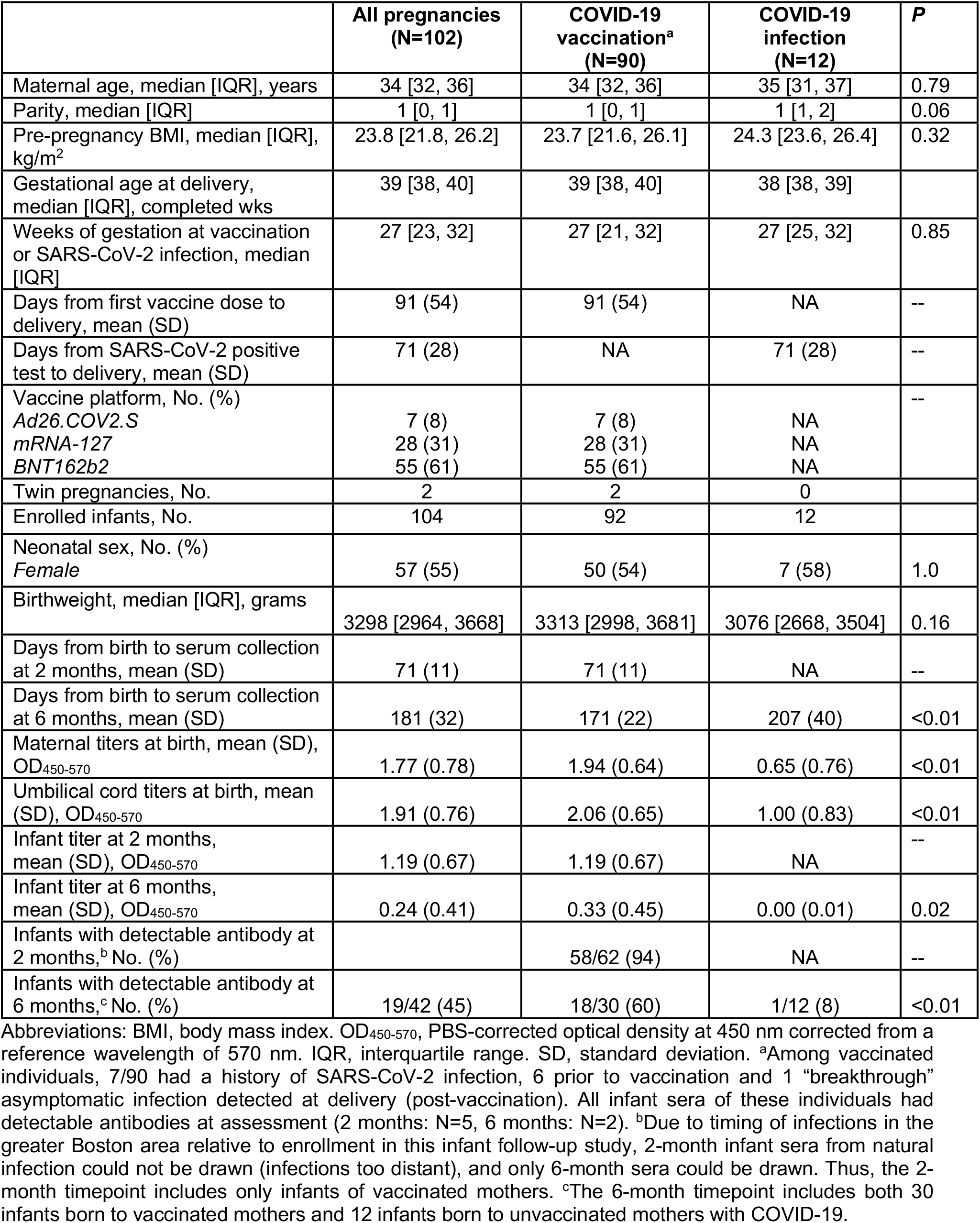

|  | All pregnancies<br>(N=102) | COVID-19<br>vaccination<sup>a</sup><br>(N=90) | COVID-19<br>infection<br>(N=12) | P |
| --- | --- | --- | --- | --- |
| Maternal age, median [IQR], years | 34 [32, 36] | 34 [32, 36] | 35 [31, 37] | 0.79 |
| Parity, median [IQR] | 1 [0, 1] | 1 [0, 1] | 1 [1, 2] | 0.06 |
| Pre-pregnancy BMI, median [IQR],<br>kg/m<sup>2</sup> | 23.8 [21.8, 26.2] | 23.7 [21.6, 26.1] | 24.3 [23.6, 26.4] | 0.32 |
| Gestational age at delivery,<br>median [IQR], completed wks | 39 [38, 40] | 39 [38, 40] | 38 [38, 39] |  |
| Weeks of gestation at vaccination<br>or SARS-CoV-2 infection, median<br>[IQR] | 27 [23, 32] | 27 [21, 32] | 27 [25, 32] | 0.85 |
| Days from first vaccine dose to<br>delivery, mean (SD) | 91 (54) | 91 (54) | NA | -- |
| Days from SARS-CoV-2 positive<br>test to delivery, mean (SD) | 71 (28) | NA | 71 (28) | -- |
| Vaccine platform, No. (%) |  |  |  | -- |
| Ad26.COV2.S | 7 (8) | 7 (8) | NA |  |
| mRNA-127 | 28 (31) | 28 (31) | NA |  |
| BNT162b2 | 55 (61) | 55 (61) | NA |  |
| Twin pregnancies, No. | 2 | 2 | 0 |  |
| Enrolled infants, No. | 104 | 92 | 12 |  |
| Neonatal sex, No. (%) |  |  |  |  |
| Female | 57 (55) | 50 (54) | 7 (58) | 1.0 |
| Birthweight, median [IQR], grams | 3298 [2964, 3668] | 3313 [2998, 3681] | 3076 [2668, 3504] | 0.16 |
| Days from birth to serum collection<br>at 2 months, mean (SD) | 71 (11) | 71 (11) | NA | -- |
| Days from birth to serum collection<br>at 6 months, mean (SD) | 181 (32) | 171 (22) | 207 (40) | <0.01 |
| Maternal titers at birth, mean (SD),<br>OD<sub>450-570</sub> | 1.77 (0.78) | 1.94 (0.64) | 0.65 (0.76) | <0.01 |
| Umbilical cord titers at birth, mean<br>(SD), OD<sub>450-570</sub> | 1.91 (0.76) | 2.06 (0.65) | 1.00 (0.83) | <0.01 |
| Infant titer at 2 months,<br>mean (SD), OD<sub>450-570</sub> | 1.19 (0.67) | 1.19 (0.67) | NA | -- |
| Infant titer at 6 months,<br>mean (SD), OD<sub>450-570</sub> | 0.24 (0.41) | 0.33 (0.45) | 0.00 (0.01) | 0.02 |
| Infants with detectable antibody at<br>2 months,<sup>b</sup> No. (%) |  | 58/62 (94) | NA | -- |
| Infants with detectable antibody at<br>6 months,<sup>c</sup> No. (%) | 19/42 (45) | 18/30 (60) | 1/12 (8) | <0.01 |

Abbreviations: BMI, body mass index. OD<sub>450-570</sub>, PBS-corrected optical density at 450 nm corrected from a reference wavelength of 570 nm. IQR, interquartile range. SD, standard deviation. <sup>a</sup>Among vaccinated individuals, 7/90 had a history of SARS-CoV-2 infection, 6 prior to vaccination and 1 “breakthrough” asymptomatic infection detected at delivery (post-vaccination). All infant sera of these individuals had detectable antibodies at assessment (2 months: N=5, 6 months: N=2). <sup>b</sup>Due to timing of infections in the greater Boston area relative to enrollment in this infant follow-up study, 2-month infant sera from natural infection could not be drawn (infections too distant), and only 6-month sera could be drawn. Thus, the 2-month timepoint includes only infants of vaccinated mothers. <sup>c</sup>The 6-month timepoint includes both 30 infants born to vaccinated mothers and 12 infants born to unvaccinated mothers with COVID-19.

**Figure 1.**
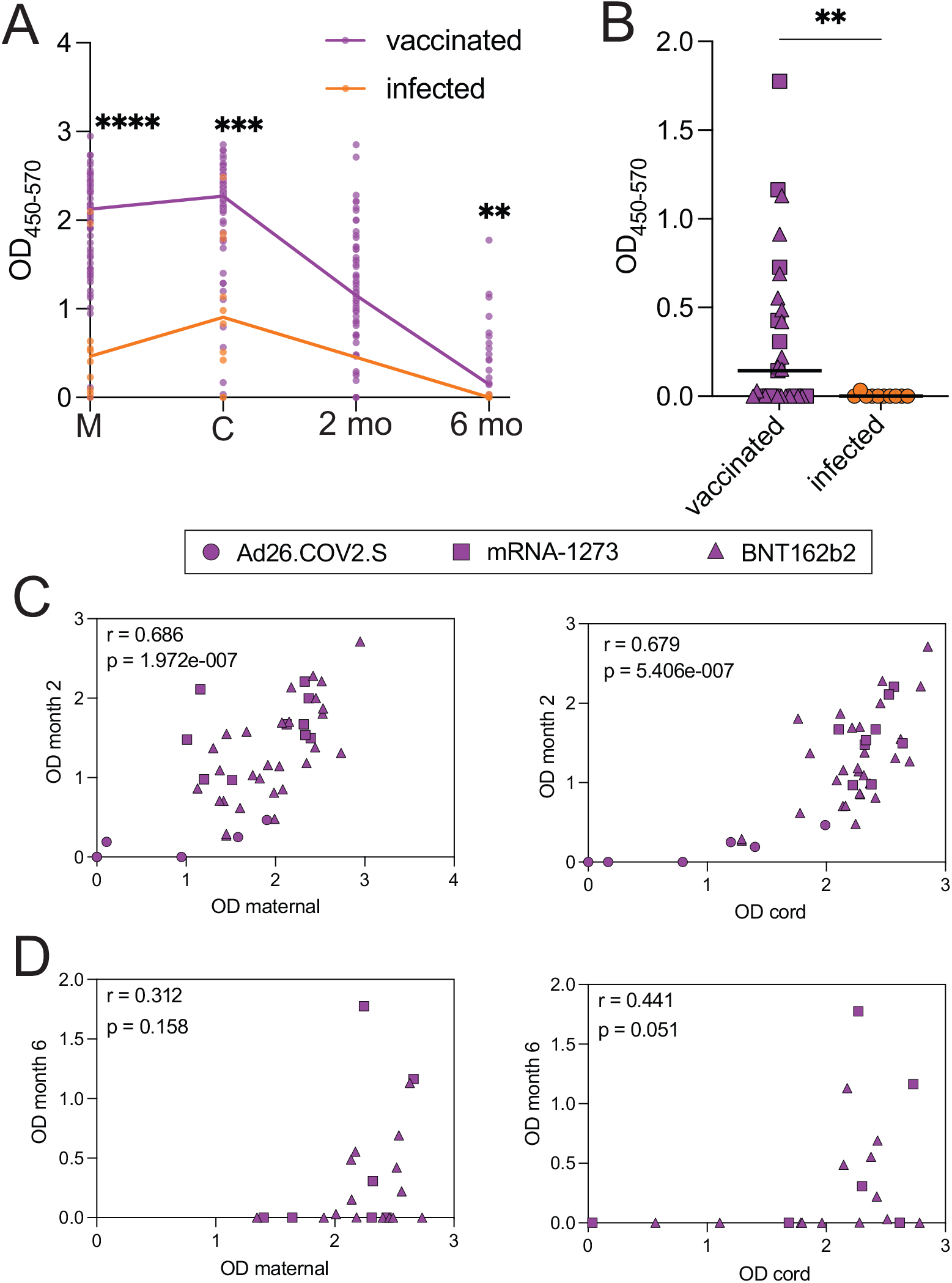
A. Plot depicting anti-S antibody titers in maternal sera at delivery (M), umbilical cord sera at delivery (C), infant sera at 2 months of age (2 mo) and infant sera at 6 months of age (6 mo) in vaccinated pregnancies (“vaccinated”, purple) and unvaccinated pregnancies infected with SARS-CoV-2 during pregnancy (“infected”, orange). Differences between groups at M, C and 6-month time points were assessed by Mann-Whitney; due to the timing of COVID-19 cases relative to the study period, infants born to women infected with COVID-19 during pregnancy were older than 2 months of age at sample collection (see Supplementary Methods). ** p < 0.01, *** p < 0.001, **** p < 0.0001. OD_450-570_, optical density (OD) at 450 nm corrected from a reference wavelength of 570 nm. B. Plot depicting infant titers at the 6-month timepoint in vaccinated (purple) and infected (orange) groups. C. Infant titers at 2 months versus maternal titers (left) and umbilical cord titers (right) in vaccinated pregnancies. Vaccine platform (Ad26.CoV2.S, mRNA-1273, BNT162b2) indicated by symbol. Spearman rho and p-value are shown for each correlation. D. Infant titers at 6 months versus maternal titers (left) and umbilical cord titers (right) in vaccinated pregnancies. Vaccine platform indicated by symbol. Spearman rho and p-value are shown for each correlation.

## Discussion

Understanding the persistence of maternal antibody in the infant is of significant clinical importance, as COVID-19 infections in this age group account for a disproportionate burden of pediatric SARS-CoV-2-associated morbidity,^5^ and COVID-19 vaccines are not currently planned for administration to infants younger than 6 months. Here we demonstrate that, consistent with prior findings by our group and others,^1,3^ vaccination resulted in higher maternal and cord titers than natural COVID-19 infection. While there is no known antibody titer at which protection from COVID-19 is assured, 60% of infants born to women vaccinated during pregnancy had detectable titers at 6 months. The finding that the majority of infants (11/12) had no detectable titer at 6 months after maternal natural infection is consistent with the limited data available about antibody persistence after natural infection,^6^ and indicates a relative deficit in protective immunity transferred to the infant compared to maternal vaccination.

These data point to the importance of immunization against SARS-CoV-2 in the first year of life, as maternally-derived immunity may only last 4-6 months. The optimal timing for vaccination in the first year of life against SARS-CoV-2 is not known; balancing maternal antibody persistence and potential vaccine interference with achieving neonatal protection remains a key consideration. The role of breastmilk antibodies in bridging infant protection while placentally-transferred antibodies decay also remains incompletely understood, but together these two forms of passive immunity will holistically inform the timing of immune vulnerability to SARS-CoV-2. These data provide further incentive for pregnant individuals to pursue COVID-19 vaccination during pregnancy to maximize protection for themselves and their infants.

## Data Availability

All data produced in the present study are available upon reasonable request to the authors.

## Authorship statement

Dr. Shook and Ms. Atyeo contributed equally to this work. Drs. Edlow, Alter and Gray contributed equally to this work.

## Acknowledgments

K.J.G. has consulted for Illumina, BillionToOne, and Aetion outside the scope of the submitted work. G.A. is the founder of Seromyx Inc. A.F. reported serving as a cofounder of and owning stock in Alba Therapeutics and serving on scientific advisory boards for NextCure and Viome outside the submitted work. All other authors report no competing interests.

