## Supplemental Methods for "Durability of anti-Spike antibodies in the infant after maternal COVID-19 vaccination"

### Supplementary Methods

#### *Study design and participant selection*

Pregnant individuals were eligible for inclusion in the prospective longitudinal pregnancy cohort (MGB IRB #2020P003538) if they were 18 or older, received one of three available COVID-19 vaccines during pregnancy or tested positive for SARS-CoV-2 by nasopharyngeal swab during pregnancy, were able to provide informed consent, and delivered at one of two Harvard-affiliated hospitals: Massachusetts General Hospital or Brigham and Women's Hospital. Pregnant individuals vaccinated between 20 and 32 weeks' gestation were targeted for enrollment, as previous studies have demonstrated superior transplacental transfer of antibodies during this window as compared with vaccination occurring closer to delivery.[4] Participants with SARS-CoV-2 infection during the same window were approached for enrollment of their infants, as second and early third trimester infection have been demonstrated to result in improved placental transfer relative to later third trimester infection. Vaccinated individuals with a history of SARS-CoV-2 infection before or during pregnancy were included in the vaccination group as the primary objective of this study was to characterize durability of infant titers after maternal vaccination in a real-world setting. Matched maternal and umbilical cord sera were collected during the delivery hospitalization. Liveborn infants of enrolled pregnant participants were enrolled after birth (MGB IRB #2020P000955).

In a cross-sectional study design, we sought to assess infant titers either at 2 months of life or at approximately 6 months of life, as these are clinically relevant time points during which pediatric vaccines may be given, and there may be theoretical concern about persistence of infant titer resulting in antibody interference with vaccine response Infants born to pregnant individuals testing positive for SARS-CoV-2 during pregnancy between 20 and 32 weeks' were enrolled as a comparator group for antibody transfer. Due to the timing of the second wave of the COVID-19 pandemic in Massachusetts (winter to early spring 2021) relative to the period of sample collection (summer and early fall 2021), we could only capture the 6-month timepoint for sample collection in these infants.

#### *Sample collection and processing*

Maternal and umbilical cord sera were collected via venipuncture during the delivery hospitalization. Infant sera were collected via microneedle-based capillary blood collection device (Seventh Sense Biosystems). Serum separator tubes were centrifuged at 1000 g for 10 min at room temperature. Serum and plasma were aliquoted into cryogenic vials and stored at -80°C.

#### *Enzyme-linked immunosorbent assay (ELISA)*

Antibodies against the SARS-CoV-2 Spike were quantified using an ELISA. ELISA plates were coated with 500 ng/mL of D614G Spike (kindly provided by Erica Saphire) and incubated for 30 minutes at room temperature. Plates were washed with washing buffer (0.05% Tween-20. 400mM NaCl, 50mM Tris, pH 8.0) and blocked with a 0.1% BSA solution for 30 minutes at room temperature. Plates were washed, and sample was added at a dilution of 1:100. Plates were incubated with sample at 37°C for 30 minutes. Plates were washed, and a horseradish peroxidase (HRP)-conjugated goat anti-human IgG antibody (Bethyl Laboratories) was added for detection of Spike-specific IgG. Plates were incubated with secondary antibody for 30 minutes at room temperature and then washed. TMB was used to develop the ELISA and sulfuric acid was used to stop the ELISA. Signal was read at 450 nm and PBS-background corrected from a reference wavelength of 570 nm.
